# Modeling shield immunity to reduce COVID-19 transmission in long-term care facilities

**DOI:** 10.1101/2021.07.16.21260657

**Authors:** Adriana Lucia-Sanz, Andreea Magalie, Rogelio Rodriguez-Gonzalez, Chung-Yin Leung, Joshua S. Weitz

**Affiliations:** School of Biological Sciences, Georgia Institute of Technology, Atlanta, GA 30332, USA; Interdisciplinary Graduate Program in Quantitative Biosciences, Georgia Institute of Technology, Atlanta, GA 30332, USA; School of Physics, Georgia Institute of Technology, Atlanta, GA 30332, USA

**Author notes:** Systems Modeling and Translational Biology, GSK, Stevenage, SG1 2NY UK. These authors contributed equally to this work (Adriana Lucia-Sanz, Andreea Magalie, Rogelio Rodriguez-Gonzalez).

## Abstract

Nursing homes and other long-term care facilities in the United States have experienced severe COVID-19 outbreaks and elevated mortality rates, often following upon the inadvertent introduction of SARS-CoV-2. Following FDA emergency use approval, widespread distribution of vaccines has resulted in rapid reduction in COVID-19 cases in vulnerable, older populations. Yet, vaccination coverage remains incomplete amongst residents and healthcare workers. As such, mitigation and prevention strategies are needed to reduce the ongoing risk of transmission and mortality amongst vulnerable, nursing home populations. One such strategy is that of ‘shield immunity’, in which recovered individuals increase their contact rates and therefore shield individuals who remain susceptible to infection. Here, we adapt recent population-scale shield immunity models to a network context. To do so, we evaluate network-based shield immunity by evaluating how restructured interactions in a bipartite network (e.g., between healthcare workers and long-term care residents) affects SARS-CoV-2 epidemic dynamics. First, we identify a series of rewiring principles that leverage viral testing, antibody testing, and vaccination information to reassign immunized healthcare workers to care for infected residents while retaining workload balance amidst an outbreak. We find a significant reduction in outbreak size when using infection and immune-based cohorting as a weekly intervention. Second, we also identify a preventative strategy using shield-immunity rewiring principles, by assigning susceptible healthcare workers to care for cohorts of immunized residents; this strategy reduces the risk that an inadvertent introduction of SARS-CoV-2 into the facility via a healthcare worker spreads to susceptible residents. Network-based epidemic modeling reveals that preventative rewiring can control the size of outbreaks at levels similar to that of isolation of infectious healthcare workers. Overall, this assessment of shield immunity provides further support for leveraging infection and immune status in network-based interventions to control and prevent the spread of COVID-19.

## Introduction

SARS-CoV-2 remains a global threat as of July 2021 with more than 33M documented cases and 600K fatalities in the US alone, and more than 187M cases and 4M fatalities worldwide. In the US, nursing homes and long-term care facilities have experienced severe outbreaks and elevated death rates [1]. Both residents and staff have been disproportionately affected by SARS-CoV-2 compared to other population groups [2, 3, 4]. Coronavirus disease (COVID-19) affects the elderly far more severely, on average, than younger individuals [5]. Besides age, other high-risk factors for COVID-19 severity in nursing homes and long-term health care facilities (which we refer to as LTCs) include co-occurring conditions, such as cardiovascular disease, chronic respiratory disease, and diabetes [6, 7]. In addition to housing vulnerable populations, nursing homes are also the site of high transmission rates owing to high density, levels of contact and challenges in infection control. This increased risk is evident in the gap between cases and fatalities in the US: as of March 19, 2021, 34% of COVID-19 fatalities were in LTCs compared to just 4% of documented cases [8].

From the outset, there have been acute challenges in preventing and responding to COVID-19 outbreaks in LTCs. As of early May 2020, thousands of LTCs across the U.S. reported cases of COVID-19 among residents and staff, given limitations to prevention policies, facility-wide testing, and support for staff [9]. Data from June 2020 reported that 71% of 13,167 US nursing homes had at least one case among residents and/or staff and 27% of facilities reported an outbreak [10]. Understaffing [10] and staff movement across facilities [11] have shown to be important factors for COVID-19 outbreaks among nursing homes. The combination of those high-risk factors, vulnerable residents sharing space and requiring prolonged and intense contact with staff seem to have been critical for the severity of the COVID-19 pandemic in LTCs [12, 13].

The increasing availability of highly effective and safe vaccines has contributed to the rapid decline in severe cases of COVID-19 amongst vulnerable individuals [14, 15]. However, vaccine coverage remains incomplete, amongst residents and especially among staff. Protection of healthcare workers (HCWs) who are at increased risk to become infected by COVID-19 [16, 17] is of paramount importance for the care of residents and might be fundamental to control ongoing and future outbreaks [18, 19]. Hence, enhanced protocols are urgently needed to combat COVID-19 transmission in nursing homes and other LTCs. Amongst non-pharmaceutical interventions, recommendations have centered on testing, cohorting and restricting movement across and within facilities. Facility-wide surveillance testing, either via antigen or molecular viral testing, provides a mechanism to identify and isolate residents as well as to reduce the risk that infected staff interact with other staff members and vulnerable residents [20]. As a complementary approach, models of staff cohorting could lead to fewer infections among HCW when staff groups work in weekly shifts [21]. Designing cohorting interventions based, in part, on immune status (rather than infection status alone) remains under-explored.

One way to leverage testing to improve infection control is to change which HCWs care for which residents based on both disease and immune status. Shield immunity represents one strategy to leverage antibody tests to reduce transmission risk, such that recovered individuals increase their contact rates, including with susceptible individuals [22]. As a result, the frequency of potential risky interactions between individuals of unknown status (including susceptible and infectious individuals) are reduced. Follow-up modeling work showed that implementation of shield immunity could retain effectiveness at population scales even with imperfect tests [23] and could help balance public health and socioeconomic outcomes [24]. Adapting a shield immunity strategy for implementation in LTCs requires specifying which HCWs care for which residents as part of a dynamic epidemic network model (*sensu* [25]). Re-assigning interactions in a bipartite network (i.e., representing interactions between HCWs and residents) may also be logistically challenging due to workload balance and other constraints.

Here, we use a network model approach to study the effectiveness of shield immunity in reducing outbreak size in LTCs. We propose an immune shielding rewiring algorithm that implements cohorting and workload assignments between HCWs and residents based on disease status. Consistent with prior work, we find that outbreak size can be reduced when immunized HCWs care for infected residents. Network simulations show that when immune shielding rewiring is implemented weekly, then shield immunity-based “rewiring” can reduce outbreaks beyond that achieved by viral testing alone. We also develop a preventative “prewiring” intervention and show that cohorting susceptible HCWs with recovered or vaccinated residents could prevent future outbreaks - because an inadvertent introduction of SARS-CoV-2 is less likely to spread when susceptible HCWs provide care for immune residents. This prewiring intervention may provide one route to decrease risks of outbreaks in partially vaccinated populations of HCWs. Overall, this network modeling study provides further evidence that identifying and leveraging disease status as a means to personalize interventions can be a critical part of efforts to control and prevent COVID-19, especially amongst vulnerable populations.

## Methods

### Summary

We simulate the spread of SARS-CoV-2 in a nursing home via a stochastic network-based model. We use an SEIR representation of disease states and a network consisting of two sets of nodes: HCWs and residents. Every individual is represented by a node which can be either susceptible S, exposed E (contracted SARS-CoV-2 but not yet infectious), infectious I, or recovered R (acquired immunity to SARS-CoV-2 and no longer infectious). We note that our I class contains both symptomatic and asymptomatic individuals. Every time step (10 minutes), individuals interact with exactly one of their neighbors with probability *P*_*contact*_ such that every individual averages *β*=*0*.*5* contacts through which infection can spread per day. Infection spreads strictly interactions between I and S individuals; newly infected individuals enter the E class. Further, at every time step exposed individuals become infectious with probability *P*_*EI*_ and infectious individuals recover with probability *P*_*IR*_. We note that a full treatment of heterogeneity in interaction intensity (e.g., between HCWs based on work category and duties as well as between residents based on room location) is beyond the scope of the present model (see Discussion for more details on potential extensions).

The proposed mitigation strategies (immune shielding, prewiring, and isolation) depend on knowledge of the infection status of individuals. We distinguish 3 possible test status states for every individual, assuming high quality tests (i.e., high sensitivity and specificity):

- PCR negative and seronegative/not vaccinated - Susceptible
- PCR positive - Infected
- PCR negative and seropositive/vaccinated - Recovered

We assume that exposed individuals are grouped with susceptible individuals given that their PCR test status would be negative. Our models make the following implicit assumptions: the disease status of individuals is obtained at the same frequency as the mitigation interventions are applied (e.g. weekly testing is required for weekly immune shielding), the disease status of an individual does not change between when testing is performed and when the intervention is applied (i.e., delays in testing are not incorporated in the model), testing is assume to be 100% accurate, and recovered individuals cannot be reinfected. In order to apply immune shielding on a weekly basis, individuals would be tested once a week and then residents are re-assigned to HCWs based on the proposed immune shielding strategy and test status. Specific details on the simulations and model assumptions are described in the sections below.

### Stochastic SEIR model

We use a frequency dependent SEIR epidemiological model on a bipartite network (i.e., where interactions occur between HCWs and residents). We choose a frequency dependent model (rather than density-dependent model) to mimic social distancing guidelines in nursing homes and because of the short length of a simulated time step. Hence, we assume that within a time step of 10 minutes, an individual is in close contact with at most one other individual irrespective of the size of the facility. Depending on contact rates, individuals need not have a close contact in a particular 10 minute window.

Nodes can change their disease status at every time step based on the following three events:

1. E →I: With probability *P*_*EI*_, an exposed E node will become infectious
2. I →R: With probability *P*_*IR*_, an infected I node will become recovered
3. S →E: With probability *P*_*contact*_, a susceptible S node will have a potentially infectious contact with a random neighbor. If that neighbor is infected I, the susceptible node becomes exposed E.

The transition probabilities per time step (*P*_*EI*_,*P*_*IR*_,*P*_*contact*_)are derived from underlying parameters, e.g., the infectious contact rate *β*=*0*.*5 day*^−*1*^, exposed to infected rate of *γ*_*E*=_*1*/*2 day*^−*1*^and recovery rate of 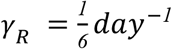 [22], as follows:

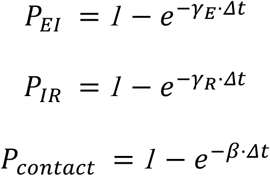

The choice of a low infectious contact rate *β*=*0*.*5 day*^-1^ reflects the use of personal protective equipment (PPE) by staff and, in some cases, by residents. The expected *R*_*0*_ for the stochastic SEIR model on a network is 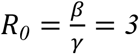 since the contact rate is independent of the degree of each node.

### Testing

To identify S or I individuals, high sensitivity and specificity PCR diagnostic tests need to be performed before applying mitigation. We assume that the PCR test correctly identifies S and I individuals (assuming high sensitivity and specificity, respectively) and will identify E individuals as S (assuming that E is of short duration and individuals in this compartment do not have sufficient viral load to generate a positive PCR result). To identify R individuals, facilities could use antibody tests, vaccination status or presume immunity within 6 months of recovery from confirmed infection. Since antibody status is maintained for an extended period of time, antibody testing could be done at a lower frequency than diagnostic testing [26].

### Network setting

We consider a bipartite network consisting of 100 healthcare workers (HCWs) and 100 residents yielding a 1:1 ratio consistent with levels of care in skilled nursing facilities. We also consider variation in this care ratio reflecting realized variation in LTCs, spanning 1:3, 1:5 and 1:10 (ratios denote HCWs:residents). Note that all synthetic bipartite networks have a mean of 1000 total edges. The choice of a bipartite network is motivated by the strict social distancing guidelines in LTCs, assuming only necessary care-centered interactions take place. We subsequently relax this assumption and allow connections between HCWs. We use two kinds of network structures: (i) random interactions between HCWs and residents; (ii) small-world social networks for interactions amongst HCWs. We construct a random bipartite network with an average degree of 10 [27], in practice this yields a binomial degree distribution with minimum degree 3 and maximum degree 20. When HCW-HCW interactions are considered, we simulate the network of interactions as a Watts-Strogatz social interaction network with average degree 10 and edge rewiring probability p = 0.02 [28].

## Mitigation strategies

### Immune shielding rewiring algorithm

We adapt a network ‘rewiring’ algorithm which provides an efficient and unbiased method to randomize connections between nodes while preserving their degree [29]. The adaptation focuses on rewiring to fulfill two key objectives (i) Minimize *I*_*Resident*_− *S*_*HCW*_ connections; (ii) Minimize *S*_*Resident*_ −*I*_*HCW*_ connections. To minimize *I*_*Resident*_ − *S*_*HCW*_ connections, we find all residents that are in the I state and all residents that are in either the R or S state. We use the notation *I*_*Resident*_ as well as *R*_*Resident*_ or *S*_*Resident*_ to refer to a resident drawn from these sets, respectively. We use a similar notation to refer to healthcare workers. Given *N*_*I*_ infected residents and *N*_*RS*_ recovered or susceptible residents, we perform the following algorithm *N*_*I*_ * *N*_*RS*_ times (S1 Fig):

1. Randomly select an *I*_*Resident*_ and a *R*_*Resident*_ or *S*_*Resident*_.
2. Find all *S*_*HCW*_ connected to the *I*_*Resident*_, but not to the *R*_*Resident*_ or *S*_*Resident*_ and all *R*_*HCW*_ or *I*_*HCW*_ connected to the *R*_*Resident*_ or *S*_*Resident*_ but not to the *I*_*Resident*_.
3. Randomly reconnect the *S*_*HCW*_ with the *R*_*Resident*_ or *S*_*Resident*_, and *R*_*HCW*_ or *I*_*HCW*_ with the *I*_*Resident*_. These reconnections are termed a ‘swap’.

At the completion of this sequence of steps, the network is rewired with the same degree for each HCW and each resident; hence the workload balance of HCWs is maintained and each resident receives the same level of care (see Fig 1B).

**Fig 1.**
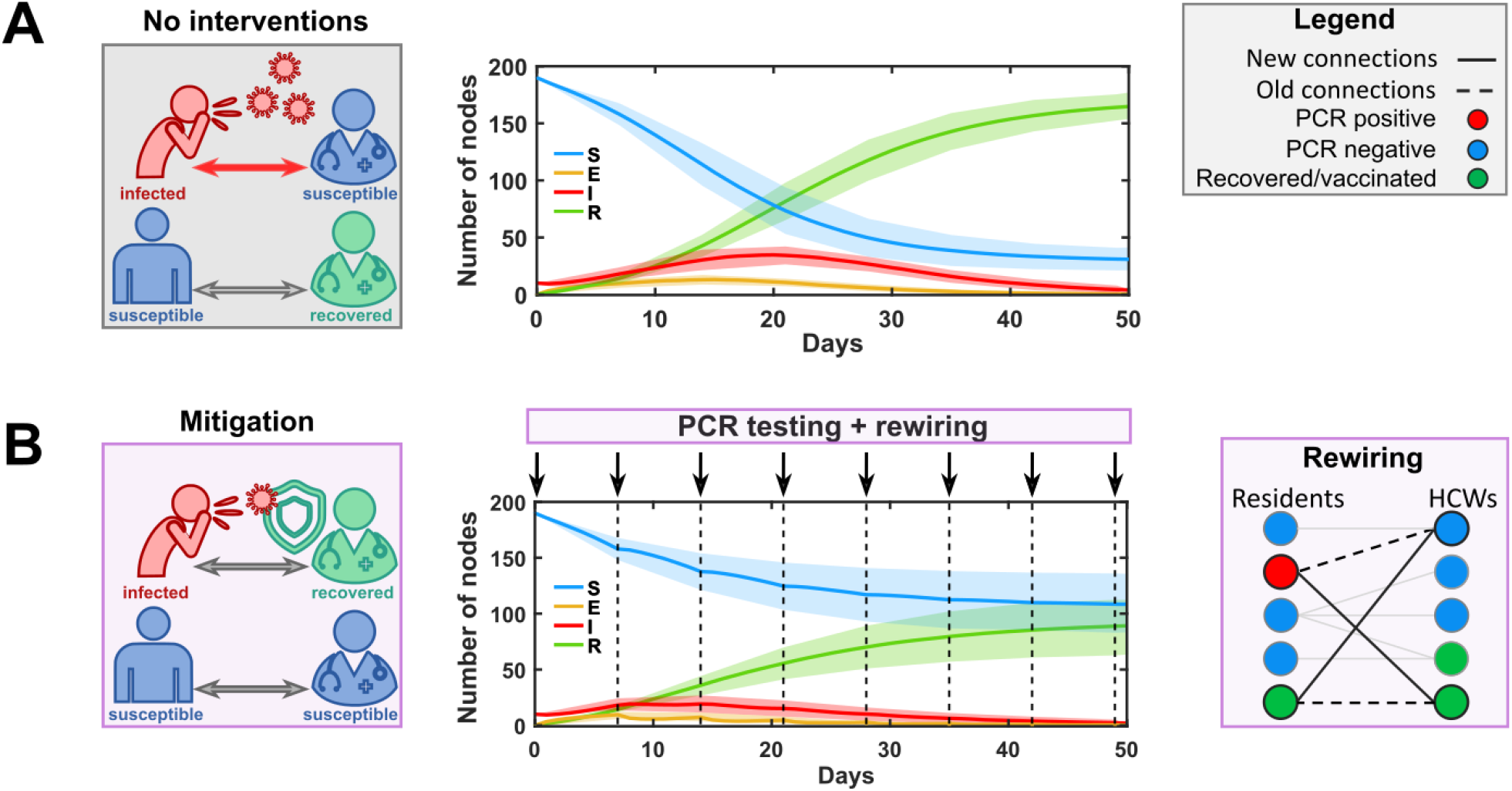
Shield immunity as a mitigation intervention in a LTC setting. Schematics (left), SEIR dynamics on a bipartite network (middle), and an example of shield immunity as a mitigation ‘rewiring’ strategy (right). SEIR dynamics show the number of nodes in S (blue), E (orange), I (red), and R (green) epidemic states. The LTC facility is represented as a bipartite network with nodes of two types: residents and HCWs. Interactions among HCWs and residents are represented as connections between nodes. Node colors show individuals PCR or immunization status as depicted in the legend. (A) Case with no interventions: we seed the epidemic with 5% of the total population (10 nodes) and simulate the outbreak over 50 days. Solid lines show the average of 500 simulation runs and shaded areas represent the standard deviation of the runs. (B) Shield immunity as a mitigation strategy: We seed the epidemic as in A. Arrows and vertical dashed lines indicate when PCR testing and rewiring are applied during the outbreak (weekly). The network shows an example of the rewiring algorithm. It deletes SI and RR (or RS) connections (dashed bolded line) and replaces them with RI and SR (or SS) connections (solid bolded line). For a complete schematic see S1 Fig.

### Prevention prewiring algorithm

We extend the rewiring algorithm to a ‘prewiring’ intervention in which there is not an outbreak (all nodes are in the S or R state). The goal of prewiring is to reconfigure interactions to minimize both the likelihood and size of an outbreak in the event of an introduced case into a LTC facility. At the network level, the prewiring algorithm minimizes the number of R-R connections while maintaining the degrees of all nodes. In effect, prewiring replaces R-R and S-S connections with R-S and S-R connections (Fig 4B). We adapt our immune shielding algorithm in the following way. We find all *R*_*Resident*_ and all *S*_*Resident*_. Given *N*_*R*_ recovered residents and *N*_*S*_ susceptible residents, we perform the following algorithm *N*_*R*_ * *N*_*S*_ times:

1. Randomly select a *R*_*Resident*_ and a *S*_*Resident*_.
2. Find all *R*_*HCW*_ connected to the *R*_*Resident*_, but not to the *S*_*Resident*_ and all *S*_*HCW*_ connected to the *S*_*Resident*_ but not to the *R*_*Resident*_.
3. Randomly reconnect *R*_*HCW*_ with the *S*_*Resident*_ and *S*_*HCW*_ with the *R*_*Resident*_.

### Isolation of infected HCWs

The isolation intervention is implemented when infectious HCWs are identified via viral testing and become “isolated” such that they do not interact with anyone until they recover from the infection. Similar to immune shielding, isolation can be implemented at different frequencies such as daily, weekly, etc. When isolated, HCWs transition to recovered (with probability *P*_*IR*_ at every time step), at which point they reconnect with their previous neighbors. Because we do not distinguish between symptomatic and asymptomatic cases, HCWs do not isolate at symptom onset but when they receive a positive PCR test.

## Numerical Simulation

The network model is implemented in MATLAB 9.7.0.1296695 (R2019b) Update 4. We run the simulation with a time step of 10 minutes and total time of 100 days. For ensemble analysis, a total of 500 simulations are run to compute the mean and standard deviations of outcomes. All outbreak simulations begin with 10 infected HCWs (10% of total HCWs) selected at random and the rest of the population susceptible, unless otherwise mentioned. We choose these initial conditions to avoid stochastic fade-out in our simulations. Prewiring based interventions assume different levels of recovered individuals as described in the Results. Code is available via https://github.com/WeitzGroup/Networks_Immune_Shielding.

## Results

### Immune shielding through rewiring infected individuals protects susceptible individuals

We evaluated the performance of the shield immunity rewiring strategy on a random bipartite network (N = 200), where half of the nodes represent residents and the other half represent HCWs. To do so, we simulated an outbreak on the network over 100 days with and without applying a dynamic rewiring strategy that leverages immune shielding on a weekly basis; resulting dynamics are shown in Fig 1B. In all cases, we focus on outbreaks with an initial size of 10, intended to evaluate the effect on interventions conditional upon epidemic liftoff. Applying the rewiring intervention weekly resulted in a 45% reduction in the epidemic peak (epidemic peak without intervention, mean = 33 infectious people, SD = 9 infectious people vs epidemic peak with weekly immune shielding intervention, mean = 18 infectious people, SD = 7 infectious people) and a 48% reduction in the final epidemic size (epidemic size without intervention, mean = 160 people, SD = 8 people vs epidemic size with immune shielding intervention, mean = 83 people, SD = 27 people). In effect, the rewiring strategy decreases the risk that infectious residents are cared for by susceptible HCWs compared to immune HCWs.

We extend our analysis to include different staffing levels consistent with 1:3, 1:5, and 1:10 HCW per resident ratios, consistent with the recommended standards for nursing homes [30]. The model predicts fewer infections when staffing levels are low in comparison to the 1:1 HCW per resident ratio (S3 Fig B). This is due to the bipartite structure we use to describe the LTC network, where residents are isolated in their rooms and can only interact with staff that follows strict social distancing guidelines. This leaves HCWs as the main source for infection propagation. Hence, reducing the number of propagators through a reduction in the HCW per resident ratio helps to reduce the overall size of the epidemic. As expected, reduced staffing levels resulted in a larger fraction of cumulative infections of HCWs in the outbreak.

### Immune shielding efficacy increases with testing frequency

Next, we evaluated the feasibility of a shield immunity rewiring strategy by assessing the impact of different testing frequencies on the final epidemic size in a network context. To do so, we simulated the SEIR epidemic model given the same bipartite network structure as described above over 100 days. We then applied the rewiring intervention described in Fig 1B (see Methods for details) at different frequencies spanning tests that occur daily, every three days, every five days, and weekly. As anticipated, an increase in testing decreased the final epidemic size (Fig 2). For example, when rewiring was applied every three days instead of every week, the mean epidemic size was 30 people (SD = 10) compared to the mean epidemic size of the scenario without intervention of 160 people (SD = 8); this corresponds to a reduction of 81% of the outbreak size. Even weekly rewiring can be an effective strategy, reducing the final outbreak size by more than 45% on average.

**Fig 2.**
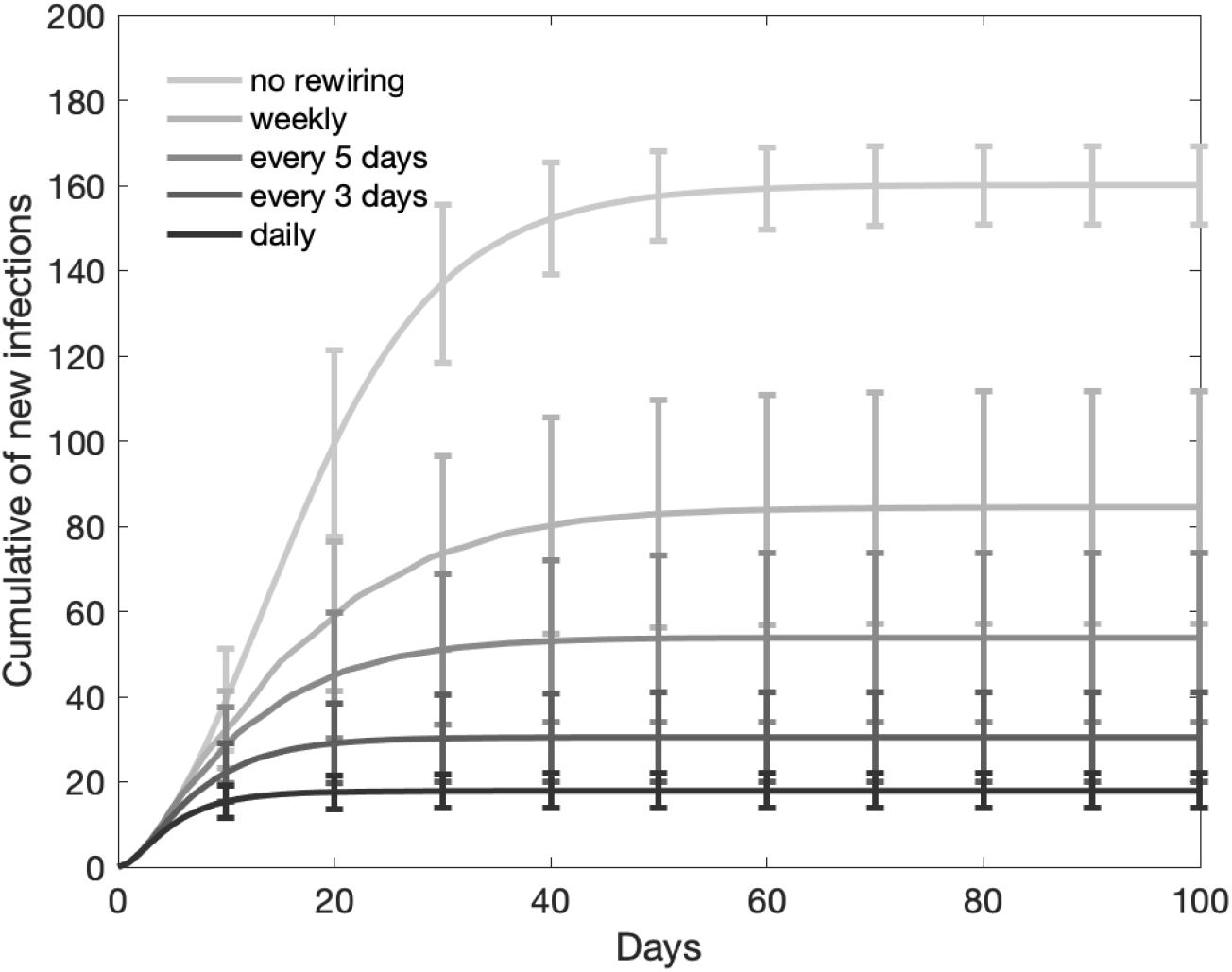
Rewiring frequency effects on outbreak size. Results of cumulative number of new infections from a SEIR model running on a random bipartite network (N = 200). The epidemic is seeded with 5% of the total population (10 nodes) infected. Lines in grayscale show the cumulative number of new infections for different rewiring frequencies (daily, every three days, every five days, weekly, and never); darker-gray lines represent more frequent rewiring schedules. New infections do not include the initial seed of 10 nodes.

### Immune shielding is potentially more effective than isolation in controlling outbreaks

We compared four scenarios to determine how beneficial immune shielding would be in a LTC facility or nursing home, in comparison to and in addition to pre-existing interventions such as isolation of infected HCWs. To do so, we ran 500 simulations of the dynamic, network model of a COVID-19 outbreak in four scenarios: (i) baseline; (ii) isolation only; (iii) shield immunity only; (iv) both isolation and shield immunity together. The baseline scenario already incorporates social distancing and other measures (e.g., partial PPE compliance) that reduces the rate of transmission. In each case, we used 500 simulations for each intervention and compared the distribution of outbreak sizes (see Fig 3A). Notably, when used on its own, shield immunity-based rewiring is more effective than isolation of HCWs: reducing the probability of having larger outbreaks (Fig 3B) and reducing the median size of outbreaks (84 people vs 122 people). We also find that combining isolation and rewiring together reduces the probability of an outbreak but does not provide a significant additional benefit in reducing outbreak sizes when outbreaks do occur. These comparative results imply that restructuring interactions is both feasible (see Fig 2) and effective (see Fig 3), even when compared to standard mitigation practice.

**Fig 3.**
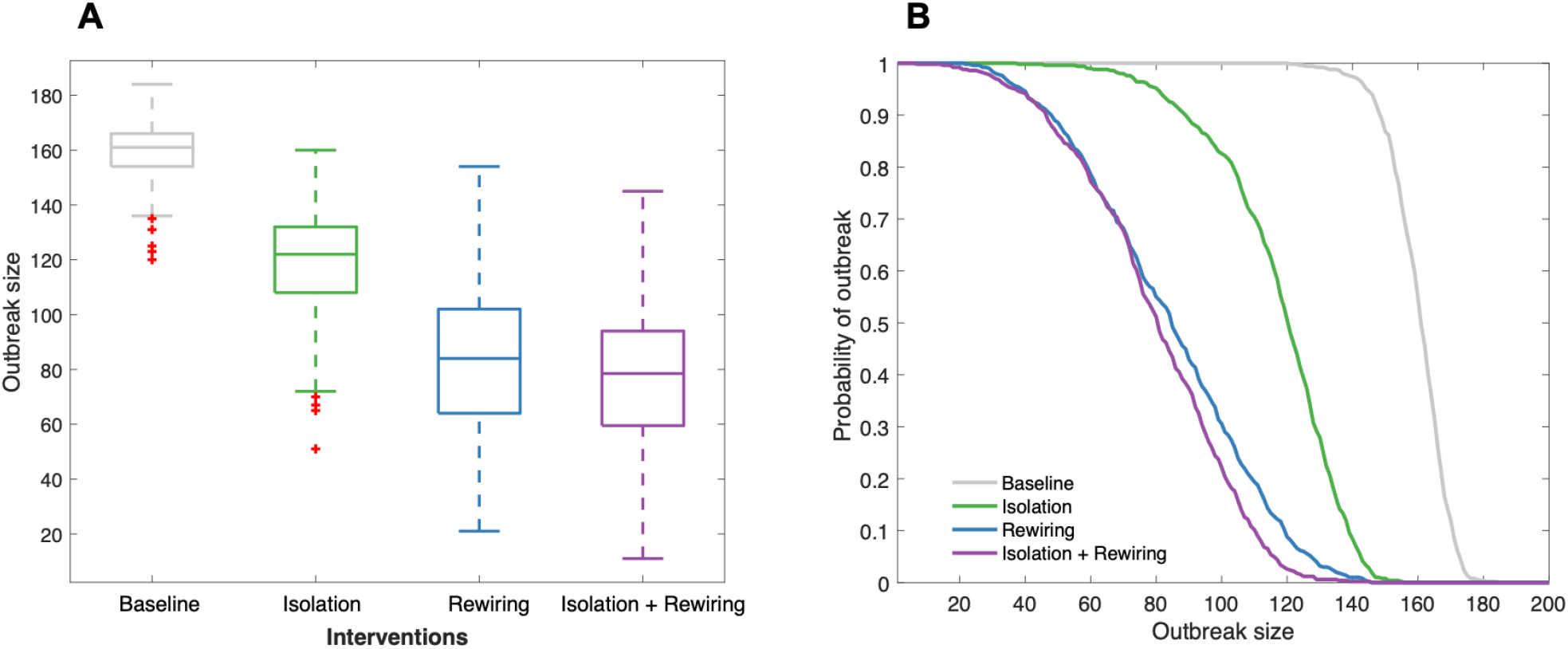
Comparison of different interventions applied on a weekly basis. (A) Distribution of the final outbreak size of 500 realizations for different interventions when we seed the epidemic with 10 infected HCWs. Boxes represent the IQR range. The mark on the box represents the median (50th percentile). Upper and lower whiskers represent 0th and 100th percentile respectively. Outliers are above or below the 1.5 the interquartile range and are shown in red + signs. (B) Probability density curves of having an outbreak of size greater or equal to the number of individuals indicated on the x-axis. All interventions are applied on a weekly basis. The outbreak size does not include the 10 nodes (5% of total population) initially used to seed the epidemic.

### Prevention of COVID-19 outbreaks in nursing homes and long-term care facilities

Thus far we have focused on evaluating the potential use of shield immunity based rewiring protocols amidst an outbreak in a nursing home or long-term care facility. But the growing rate of population immunity via recovery from prior infections and, critically, from increasing vaccination coverage suggests that it may be possible to prewire interactions to reduce the chance and size of an outbreak before outbreaks are detected. To do so, we propose a prewiring intervention that preferentially connects immune individuals with susceptible individuals to maximize immune shielding (see Methods, prewiring for details). We first compare SEIR dynamics on bipartite networks with and without applying prewiring. We simulate a second outbreak with 1 infected HCW and 30% immunized individuals in the LTC (Fig 4). We observed a reduction in the epidemic size of 44% (epidemic size without intervention, mean = 34 individuals, SD = 40 individuals vs. epidemic size with prewiring, mean = 19 individuals, SD = 27 individuals) due to prewiring. To further compare this preventive intervention with the baseline case, we calculated total number infections when we seed the epidemic with 1 infected HCW and 20, 40, 60, 80 and 100 immunized individuals: including HCW and residents (10%, 20%, 30%, 40% and 50% of the LTC). We also calculated the probability density of an outbreak given the above conditions.

**Fig 4.**
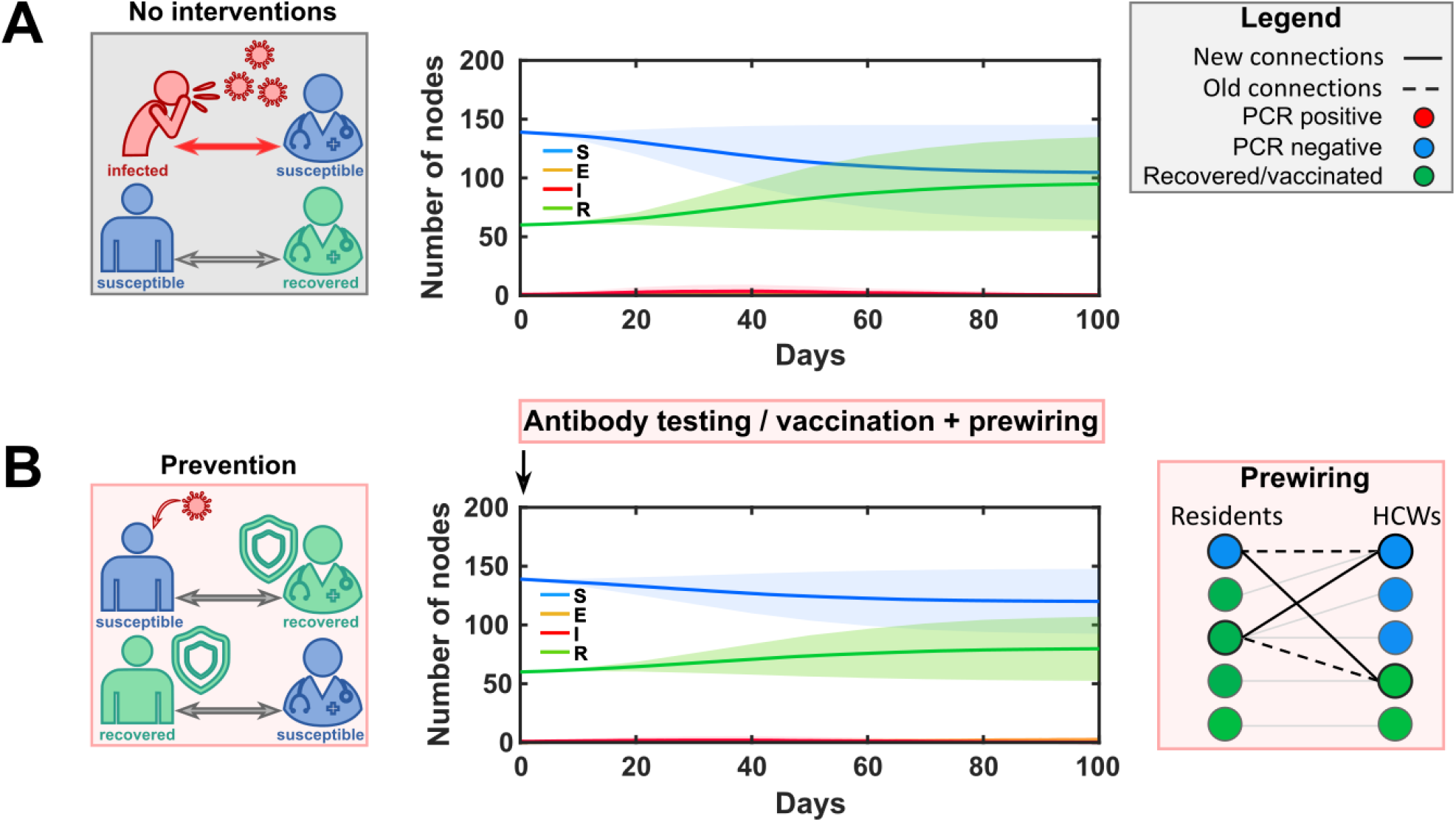
Shield immunity as a preventive intervention in a LTC setting. Schematics (left), SEIR dynamics on a bipartite network (middle), and an example of shield immunity as a preventive ‘prewiring’ strategy (right). SEIR dynamics show the number of nodes in S (blue), E (orange), I (red), and R (green) epidemic states. A second outbreak initiates with 1 infected HCW and 60 immunized (recovered/vaccinated) individuals (30% of the LTC). We simulate the epidemic over 100 days. Solid lines show the average of 500 simulation runs and shaded areas represent the standard deviation of the runs. The LTC facility is represented as a bipartite network with nodes of two types: residents and HCWs. Interactions among HCWs and residents are represented as connections between nodes. Node colors show individuals PCR or immunization status as depicted in the legend. (A) Case with no interventions. (B) Shield immunity as a prevention strategy: The arrow indicates prewiring is applied only before the outbreak starts. Prewiring rewires SS connections (dashed bold lines) and replaces them with SR connections (bold lines).

We find that preventive immune shielding significantly reduces outbreak size when immunity levels exceed 20% (Fig 5). However, prewiring interventions do not significantly reduce outbreak size when immunity levels exceed 50%; note that in such cases the outbreak size is low, even for the baseline case, in part because of the effect of preexisting susceptible depletion on disease transmission. We further compare the prewiring strategy with targeted interventions, i.e., isolation and rewiring. We show that when the immunized fraction of individuals is low (20% or less), targeted interventions (with weekly surveillance testing) are necessary to reduce the probability and size of the outbreak (S2A Fig). However, when the immunized fraction exceeds 35%, we find that prewiring intervention is as efficient as isolating infected HCWs (S2B Fig). Hence, there is an intermediate range of preexisting immunity (through natural infection and/or vaccination) in which prewiring interventions may help to reduce outbreak size in partially vulnerable populations.

**Fig 5.**
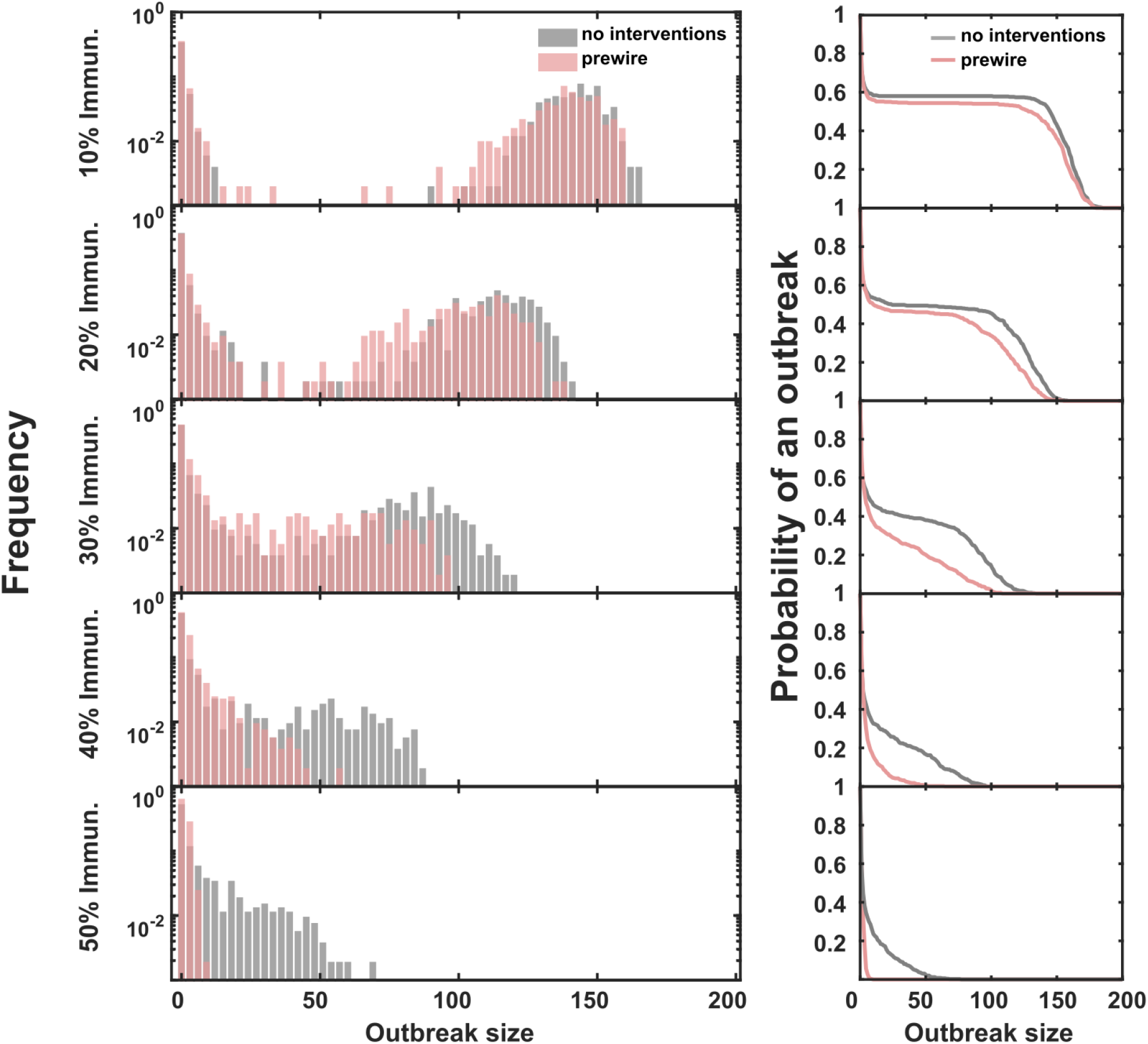
Outbreak size distributions and probability of an outbreak depending on the immunization level. Distributions of the total infected and probability densities of an outbreak for 10%, 20%, 30%, 40% and 50% of immunized individuals in the LTC when no interventions (grey) and a preventive immune shielding (prewiring, pink) strategy is applied before the outbreak starts. The epidemic initiates with 1 infected HCW. We simulate the epidemic over 100 days and perform 500 simulation runs. A two-sample Kolmogorov-Smirnov test was performed to look for a statistical significant difference of outbreak distributions with and without prewiring. P-values for 10%, 20%, 30%, 40% and 50% of immunized individuals are: 0.01994, 1.923e-05, 2.544e-11, 3.48e-10, and 2.2e-16. The distributional differences are associated with statistically significant differences in mean outbreak sizes for all but the 10% case, as quantified by a one-sided t-test with 99% confidence interval; P-values for 10%, 20%, 30%, 40% and 50% of immunized individuals are: 0.056, 0.0043, 6.5e-12, 6.9e-21, and 1.5e-24.

### Generalized prevention of COVID-19 outbreaks given staff-staff interactions

Thus far we have focused our dynamical model on risk of infection between HCWs and residents. However, previous studies have estimated that ∼50% of nursing home cases are attributable to cross-facility staff movement, hence attention to highly connected nursing facilities is warranted [11]. We followed a parsimonious scenario for a nursing home setting; the model disallows visits by family members, residents are presumably isolated in their rooms, and HCW are recommended to practice a strict social distancing protocol. Deviations of this scenario allowing HCWs to interact, even when they work in small groups or work shifts, will decrease the efficacy of immune shielding. Hence, we extended our model to include the potential for interactions between HCWs by allowing interaction links within HCWs in addition to the links between HCWs and residents. To do so, we augmented the bipartite network interactions with a small-world network representation of HCW interactions (see Methods). In S3A Fig, we show that including additional flexibility of interactions leads to an increase in cases and may require increasing the frequency of rewiring to control outbreaks and/or the inclusion of multiple prewiring steps to prevent outbreaks. In addition, we show that our findings are also robust to the modification of the ratio of HCWs to residents. S3B Fig shows that shield immunity based rewiring continues to be effective even while decreasing the HCW:resident ratio from 1:1 to 1:5. However, we note that there is a latent impact of such decreases, implying that lower ratios may actually improve infection control even in the absence of other measures given the essential role of staff in (unwittingly) mobilizing infection in a facility. Further work will be necessary to evaluate the balance between patient care, infection control, and staffing levels.

## Discussion

We developed a network-based cohorting intervention that leverages both disease status and recovery/immunization status to reduce and prevent outbreaks in nursing homes. Using a network-based intervention, we find that cohorting the care of infected residents with immunized HCWs (either via natural infection or vaccination) can significantly reduce the size of an outbreak. In doing so, the network intervention model extends prior modeling efforts to establish the benefits of antibody testing as part of a ‘shield immunity’ mitigation [22]. Using the network-based modeling framework, we also show that shield immunity principles can be applied as a preventative measure in advance of an outbreak via a prewiring step in which susceptible HCWs provide cohorted care for immune residents. This prewiring step helps to reduce the frequency and severity of outbreaks by reducing the risk that an inadvertent introduction of SARS-CoV-2 into a facility via a potentially asymptomatic HCW spreads to vulnerable residents (and then to susceptible staff). The use of weekly testing and rewiring to control an outbreak and/or the use of a prewiring step suggests that network-based cohorting interventions are likely feasible given partial population immunity.

Many nursing homes have experienced large-scale outbreaks since the onset of the SARS-CoV-2 pandemic. Despite the disproportionate impacts of severe illness in nursing homes, communities of residents and health care providers remain partially immunized, suggesting that future outbreaks of COVID-19 are expected to recur in vulnerable populations. As of June 15, 2021, approximately 50% of the eligible US population was fully vaccinated (CDC, https://covid.cdc.gov/covid-data-tracker/#vaccinations). In addition, it is likely that approximately one-third of individuals in the US have been infected and recovered from SARS-CoV-2 (CDC, https://www.cdc.gov/coronavirus/2019-ncov/cases-updates/burden.html). Immunity via natural infection and vaccination is expected to last one year or longer, although estimating reinfection risk due to variants remains an ongoing challenge. At present, best practices to prevent outbreaks in nursing homes include a combination of practices including the use of PPE, support for staff, as well as viral surveillance testing of staff and residents [31, 32, 33]. We note that our findings contrast with prior suggestions to cohort susceptible HCWs (in PPE) with infectious residents while having recovered HCWs not wear PPE when dealing with other residents [34]. We note that PPE alone is not 100% effective and that there is no longer the same constraint on the availability of PPE supplies. We caution that intentionally cohorting susceptible HCWs with infectious residents (when immunized HCWs are available) could lead to elevated risks of spread.

The present analysis leverages viral testing to inform network-based mitigation. The testing strategy leverages a high-quality viral test (analogous to a PCR test), hence real-world implementation will require considerations of trade-offs between test rate, turnaround speed, and accuracy. As is apparent, knowing both the disease and immunization status of individuals can inform shield immunity interventions. Hence, our findings suggest the value of considering large-scale antibody testing of HCW staff to inform immunity-based cohorting to reduce transmission risk, particularly in the absence of full FDA approval for vaccines (or in contexts in which mandates are not feasible, are not permitted, or are otherwise impractical). The use of both viral and antibody tests combined with vaccination mandates or surveys for vaccination status could help inform care schedules to reduce the risk of transmission of SARS-CoV-2 in nursing homes. We recognize that contact patterns in LTCs are heterogeneous and that translating the present network-based model will require inclusion of site-specific constraints.

Indeed, our network-based intervention model comes with caveats. Our focus on interventions to reduce risk of SARS-CoV-2 does not consider risks for other infections like influenza, norovirus, and antibiotic resistant pathogens, particularly if the infection risk factors for these other diseases do not coincide with those associated with SARS-CoV-2. In addition, network-based interventions require changes in staff care and availability, exploration of feasibility will require extending the current framework to reflect constraints in staff expertise, numbers, and supply. Moreover, we have assumed that recovered individuals and vaccinated individuals have protective immunity from onward transmission over the period of the epidemic outbreak (here modeled as 100 days). The duration of effective immunity has been estimated to be at least 6-8 months [35]; however, the rise of variants and heterogeneous variation in immune protection reinforce the need to use PPE in nursing home facilities until levels are substantially reduced (in contrast to some proposals that recovered individuals not use PPE, perhaps because of a concern on levels of PPE availability [34]). Applications of dynamic rewiring informed by immune shielding concepts should be evaluated and adjusted in light of the ongoing spread of variants of concern.

In summary, we have developed a network-based approach to cohort both residents and healthcare workers in light of their infection and immune status as a means to reduce the risk of active transmission or the future risk associated with the inadvertent introduction of SARS-CoV-2 into a vulnerable population. In doing so, this study reinforces a persistently under-explored opportunity: to use testing at scale as a form of mitigation rather than a passive indicator of the status of an outbreak. Here, viral testing and assessment of immune status (whether through antibody testing or via vaccination status) are combined to inform the active ‘rewiring’ or preventative ‘prewiring’ of resident to healthcare worker interactions with a central goal: reducing the size of outbreaks. With the increasing but still partial coverage of vaccines, the present study advances the goal of informing behavioral strategies to reduce the disproportionate impact of severe illness and SARS-CoV-2 associated fatalities in vulnerable, elderly populations.

## Data Availability

Simulation code is written in MATLAB R2019b and is available for download at https://github.com/WeitzGroup/Networks_Immune_Shielding

## Acknowledgements

We thank Jonathan Dushoff, Sang Woo Park, Scott Fridkin, Ben Lopman, Carly Adams, and Weitz group members for feedback on the manuscript.

## Author contributions

AL, AM, RR, CYL, JSW designed methods; AL, AM, RR, CYL developed simulations; AL, AM, RR, CYL implemented simulations; AL, AM, RR, JSW analyzed model results; AL, AM, RR, JSW wrote the paper; CYL and JSW designed the overall project.

## Supporting figures

**S1 Fig.**
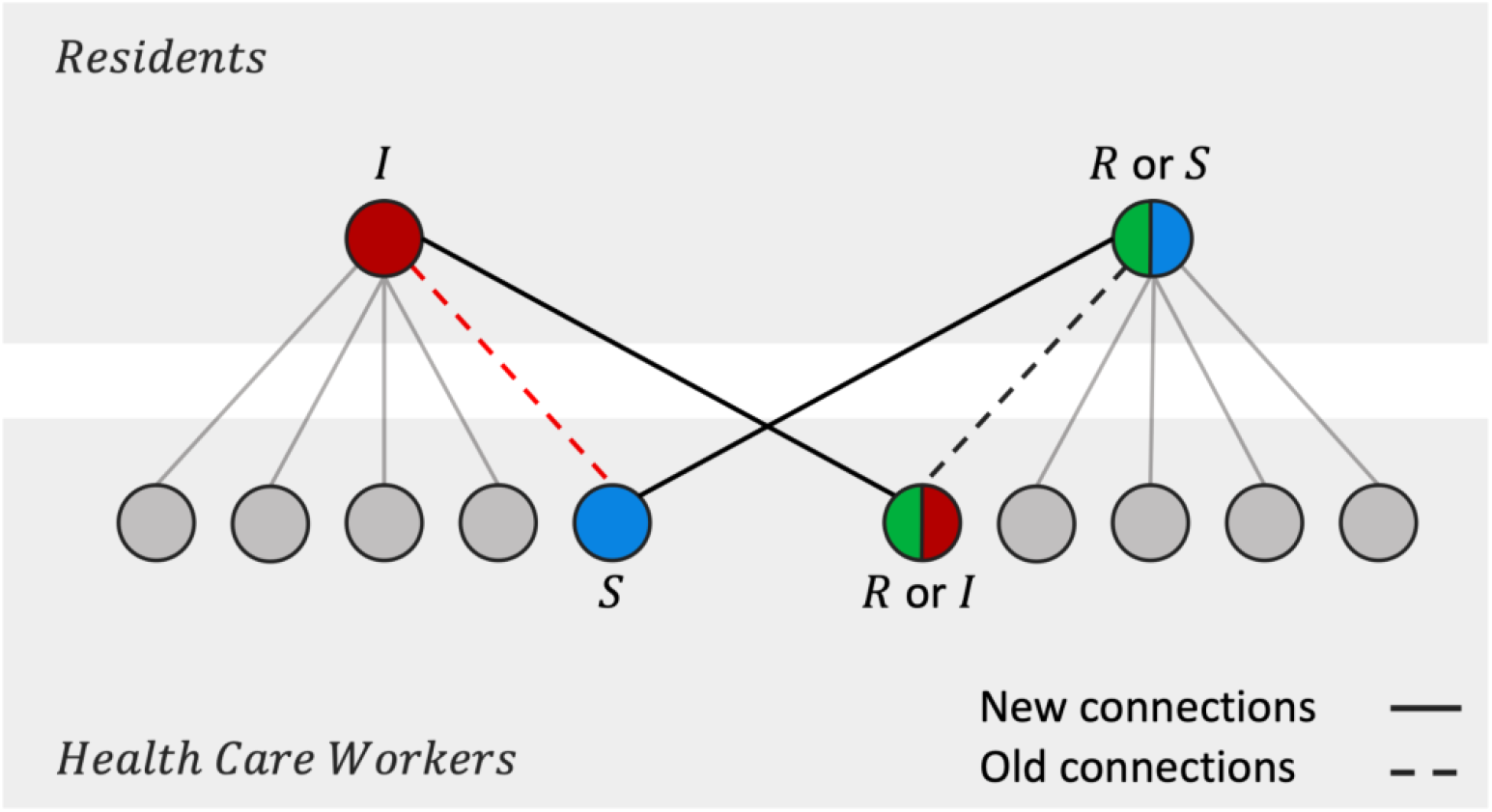
Schematic of immune shielding rewiring algorithm. Only the first part of the algorithm described in the Methods section is shown. The second part is analogous, but the labels *Residents* and *Health Care Workers* from the grey boxes would be swapped. Susceptible, infected and recovered nodes are shown in blue, red respectively green. Grey nodes have an unspecified disease status irrelevant for the rewiring mechanism. The top and bottom grey areas represent residents, respectively healthcare workers. Connections between I and S (dashed line) are replaced with connections between I and R or I (bolded line).

**S2 Fig.**
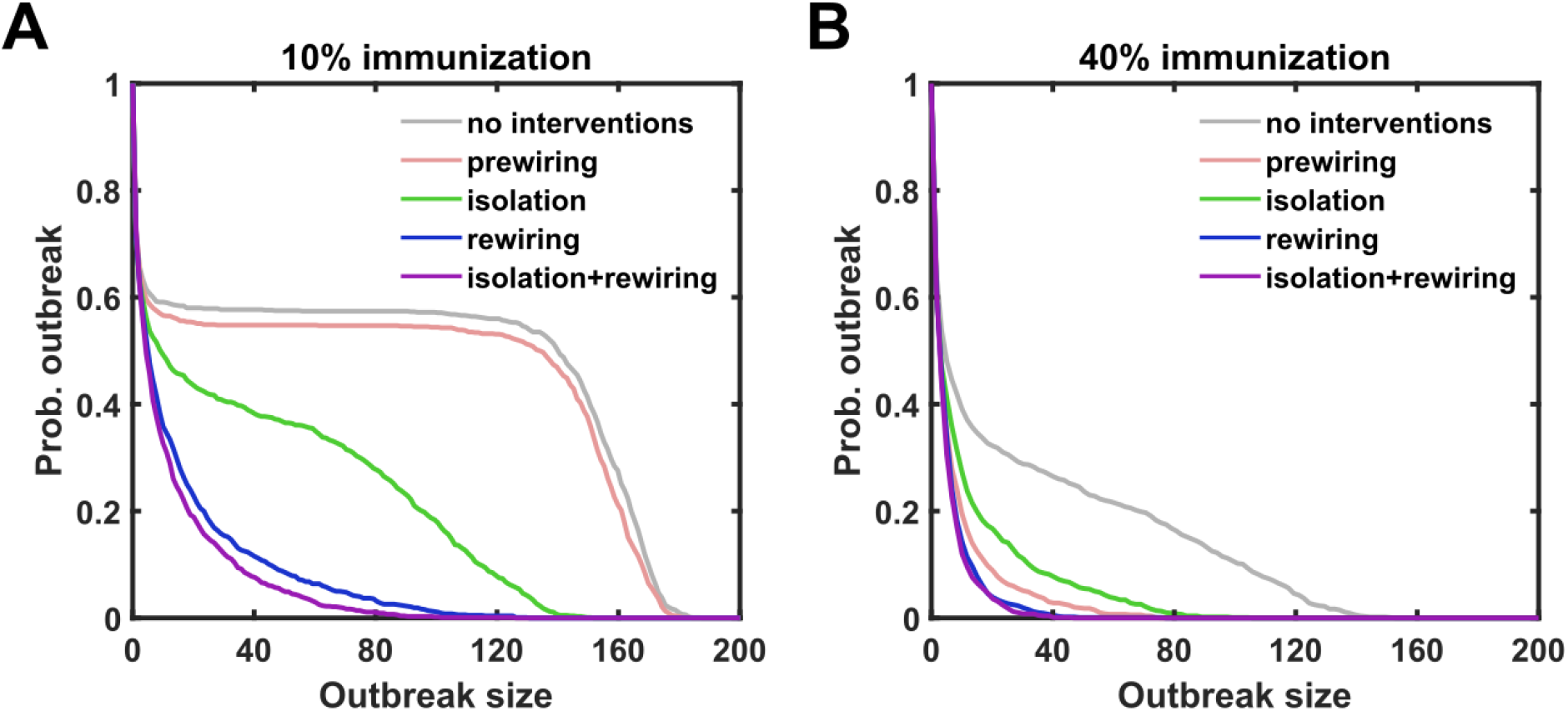
Probability density of an outbreak. Probability density curves of an outbreak of size equal or greater than number of individuals indicated in the x-axis (A) when there is a 10% immunization at the beginning of the outbreak (B) when there is 40% immunization at the beginning of the outbreak. (Grey). Case with no interventions. (Pink) Prewiring: Preventive intervention through a prewiring of the LTC where immunized residents are preferentially interacting with susceptible HCW and vice versa. (Green) Isolation: Intervention where staff is weekly tested and seropositive HCW isolate. (Blue) Weekly rewiring: Intervention of immune shielding of susceptible individuals in the facility through rewiring risky interactions of infected individuals with recovered/immunized individuals. This intervention is applied with weekly testing serostatus of the whole facility. (Purple) Isolation+rewiring: intervention that combines previous two interventions. Weekly testing of the whole facility. Seropositive HCW isolate and infected residents preferentially interact with immunized staff.

**S3 Fig.**
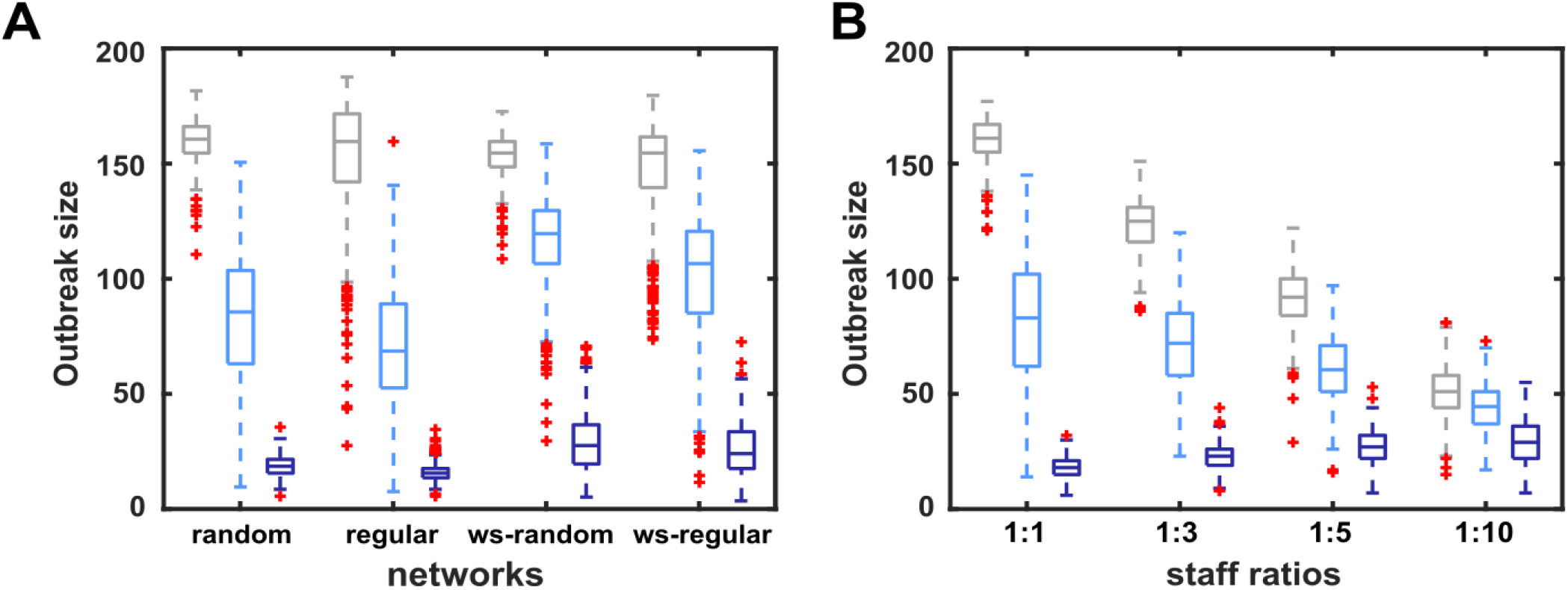
Network structure and staff ratio effects on final outbreak sizes. Final outbreak size of 500 runs when the outbreak starts with 10% of infected HCWs measured in a LTC network setting of the same size (200 individuals) and average number of connections (average degree ∼10). The connections between HCWs and residents in all LTC network settings are strictly bipartite. Grey: No interventions. Light blue: weekly rewiring. Dark blue: daily rewiring (A) Outbreak sizes for different network structures (random, regular, Watts-Strogatz random, Watts-Strogatz regular). WS-random and WS-regular networks allow connections between HCWs; regular and random networks do not allow connections between HCWs. (B) Outbreak sizes for different HCW:resident ratios in a random network LTC. Grey: No interventions. Light blue: weekly rewiring. Dark blue: daily rewiring. See Methods for details on the network structures.

